# Thalamus normalisation improves detectability of hypoperfusion via arterial spin labelling in an Alzheimer’s disease cohort

**DOI:** 10.1101/2024.08.13.24311671

**Authors:** Logan X. Zhang, Thomas F. Kirk, Martin S. Craig, Michael A. Chappell

## Abstract

Data normalisation is an important approach to reduce inter-subject variability in group studies, but care must be taken when choosing a normalisation strategy to not introduce further confounds or artefacts into the data. Normalisation of arterial spin labelling perfusion measurements remains challenging, especially in the context of Alzheimer’s disease, where there may be global hypoperfusion present. We propose that using the thalamus as a reference region for normalisation could improve the detectability of hypoperfusion in Alzheimer’s disease and alleviate the pseudo-hyperperfusion artefacts caused by the commonly-used strategy of normalisation using global mean perfusion. Evaluation on an Alzheimer’s disease dataset found this strategy was able to reduce coefficient of variation in perfusion measurements by around 60% and yield increases in statistical power of comparisons against healthy controls.

## Introduction

Alzheimer’s Disease (AD) is a neurodegenerative disorder characterised by cognitive decline and memory impairment. The pursuit of accessible and robust biomarkers for early-stage AD detection remains an active research question. Arterial spin labelling (ASL) MRI is a non-invasive physiological imaging technique that measures cerebral blood flow (CBF) [1], the disruption of which has been associated with cognitive decline in AD, and precedes morphological changes detected by structural MRI [2]. The characteristic pattern of CBF abnormality in AD subjects compared to healthy controls is hypoperfusion in the parietal lobe, precuneus and cingulate; some studies report hyperperfusion in the putamen, hippocampus, caudate and amygdala, which has been hypothesised to represent a compensatory mechanism in early-stage disease [3], [4], [5].

A confounding factor in the use of ASL to detect AD is that CBF varies due to both natural and technical factors, for example demography (females typically have higher CBF than males; CBF decreases with age [6]) or choice of scanner (differences in vendor sequence design and acquisition that affect measurement). If variability due to these non-disease factors exceeds the expected size of a disease-induced perfusion change, then it will not be possible to reliably detect disease in individuals. Normalisation of physiological activity to an area that is least affected by a disease of interest is routinely used to reduce non-disease sources of variability in neuroimaging studies [7].

A commonly-used normalisation strategy for ASL is division by a global mean CBF reference value, either to perform a simple scaling of voxel-wise CBF, or to derive a *Z*-score map [8], [9]. The assumption of this method, and indeed a desirable property of any normalisation strategy, is that the reference value should be unaffected by disease. For AD, this is a problematic assumption because both global and regional hypoperfusion are often observed, which will reduce the global mean CBF reference value [10].

Prior work applying fluorodeoxyglucose (FDG-) PET imaging to AD demonstrates the consequences of violating this assumption (metabolism, measured by FDG-PET, is closely coupled to CBF). If the reference value used for normalisation is decreased by disease compared to healthy individuals, two issues result: firstly, pseudo-hypermetabolism is observed in the regions of brain least affected by disease [11]. Secondly, the size of the change in disease-affected regions is reduced compared to healthy subjects. This is because division by an artificially small reference value inflates the normalised value. In an unaffected region, the normalised parameter appears larger than the corresponding value in a healthy subject; in a disease-affected region, the normalised parameter appears similar to the value in a healthy subject, thus reducing the contrast between different cohorts.

In FDG-PET, normalisation by the cerebellum or using ‘most-spared-region’ techniques have been successful in improving the discrimination of mild cognitive impairment from healthy aging [11], [12]. The rationale for cerebellum normalisation is that it is not substantially affected by AD; for most-spared-region techniques, statistical tests between healthy and disease groups are used to identify a region unaffected by disease. An important drawback of most-spared-region techniques is that the region must be derived for each new dataset and cannot be assumed to generalise to other datasets, particularly if a different scanner or acquisition has been used. This limits their applicability in clinical practice compared to using an anatomically-defined reference region such as the cerebellum.

For ASL, normalisation using the cerebellum is an impractical strategy as it is not always included in the acquisition field of view. Though the putamen has been used as an alternative, it may be challenging to obtain consistent reference values from this region given its small size, the low spatial resolution and low signal-to-noise ratio (SNR) of ASL data [13]. The objective of this work was therefore to identify an alternative structure for normalisation of ASL data, meeting the requirements of being unaffected by disease and sufficiently large such that it can reliably be measured using ASL. We have identified the thalamus as meeting these requirements and here evaluate its application to normalisation of an AD dataset.

## Methods and data

A previously-acquired dataset of 32 healthy controls (HC) and 23 AD subjects with clinical diagnosis was used [8]. The ethics committee of the Clinical Medical College, Yangzhou University granted approval for the study for which this data was originally acquired (decision J-2014066). T1-weighted structural images and a single-delay PCASL image were acquired on a GE Discovery MR750 3.0T scanner with an 8-channel phased-array head coil. The parameters for the T1 image were TR = 12ms, TE = 5.1ms, FOV = 256×256mm, matrix size = 256×256, 1mm slice thickness, and 0mm slice spacing. The parameters for the PCASL image were TR = 4844ms, TE = 5.1ms, PLD = 2025ms, label duration = 1450ms, FOV = 240×240mm, matrix size = 64×64 resulting in voxel size = 3.75×3.75×4mm^3^, and number of excitations (NEX) = 3; a 3D readout was used; an M0 image was acquired with TR = 4844ms.

T1 structural images were processed using FreeSurfer 6.0.1 to obtain cortical parcellations and subcortical segmentations [14]. ASL images were processed using a variant of the BASIL pipeline implementing stochastic variational Bayesian inference for perfusion estimation [15], [16]. ASL image deblurring to correct for the effects of 3D readout was performed using an in-house technique. CBF maps were calibrated to units of ml/100g/min using voxel-wise division by the M0 image. CBF measures were reported against the DKT segmentation produced by FreeSurfer [17].

Three normalisation strategies were investigated: no normalisation (calibrated CBF units alone), normalisation by mean CBF (calculated across all brain voxels within the FreeSurfer derived brain mask, excluding cerebellum and registered into ASL space), and normalisation by mean CBF calculated across the left/right thalamus. Three groups of segmentation ROIs were selected for comparison of the normalisation strategies. The first group are commonly reported to exhibit hypo-perfusion in the literature, including the precuneus, posterior cingulate gyrus, superior and inferior parietal lobes. The second group are reported to exhibit hyper-perfusion in early AD, which could either be due to a disease compensatory mechanism or pseudo-hyperperfusion artefacts. This includes the subcortical structures of the hippocampus, the caudate, and the putamen. The third group is along the central gyrus which are hypothesised to be relatively unaffected by AD, including the precentral, the postcentral and the paracentral gyri.

The performance of each strategy was evaluated by calculating the coefficient of variation (CoV, indicative of inter-subject variability) within the HC group, and by calculating paired *t*-tests between the cohorts (not assuming equal variance between the two samples). Larger *t*-values were interpreted to indicate a greater disease effect of interest. In all analyses, the threshold of statistical significance was set at *p* < 0.05.

## Results

Figure 1 shows distributions of mean CBF and thalamus CBF across the HC and AD groups. These were the reference values used for normalising CBF in the later analyses. Mean CBF was significantly different between the two groups (HC 80.93 ml/100g/min vs. AD 66.28 ml/100g/min, *p* < 0.05), whereas thalamus CBF was not significantly different (HC 75.87 ml/100g/min vs. AD 71.31 ml/100g/min, *p* =0.053).

**Figure 1:**
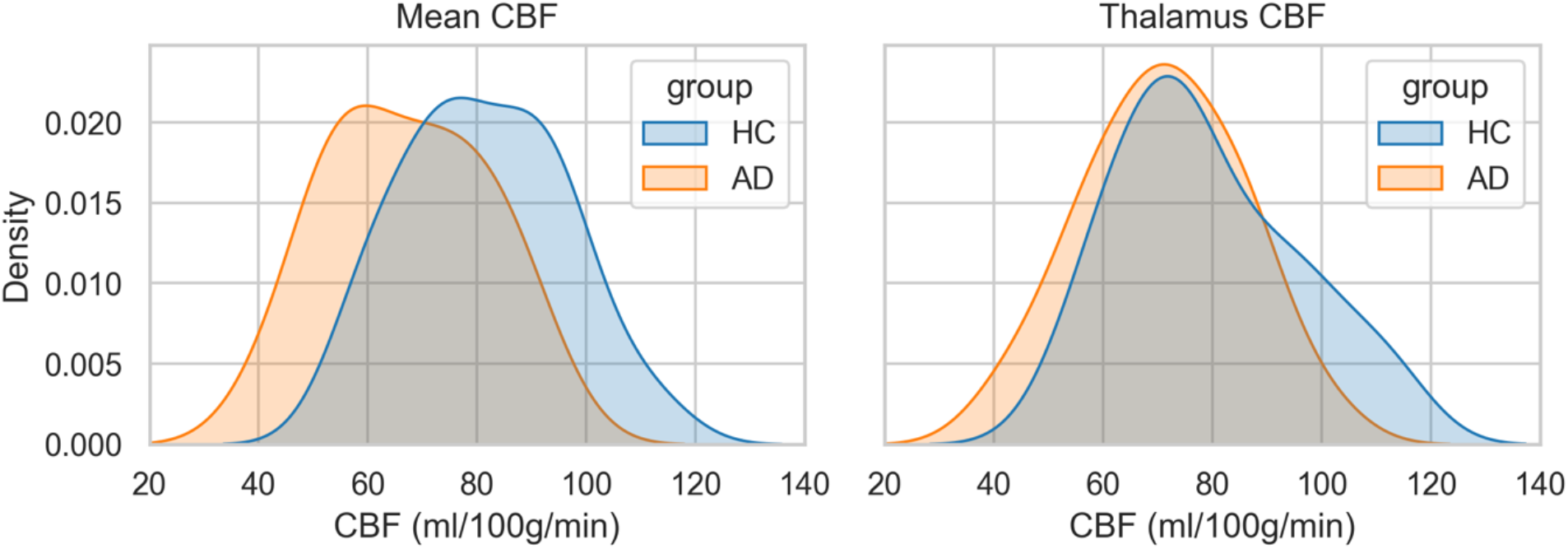
probability density plots of reference values used for normalisation within each group. Left: mean CBF was significantly different between the HC and AD groups. Right: thalamus CBF was not significantly different between the two groups.

Figure 2 shows the coefficients of variation of median CBF in each ROI, averaged across the HC subjects. Both normalisation by mean CBF and by thalamus greatly reduced the CoV compared to no normalisation, though the normalisation by mean CBF typically had a slightly lower CoV than normalisation by thalamus.

**Figure 2:**
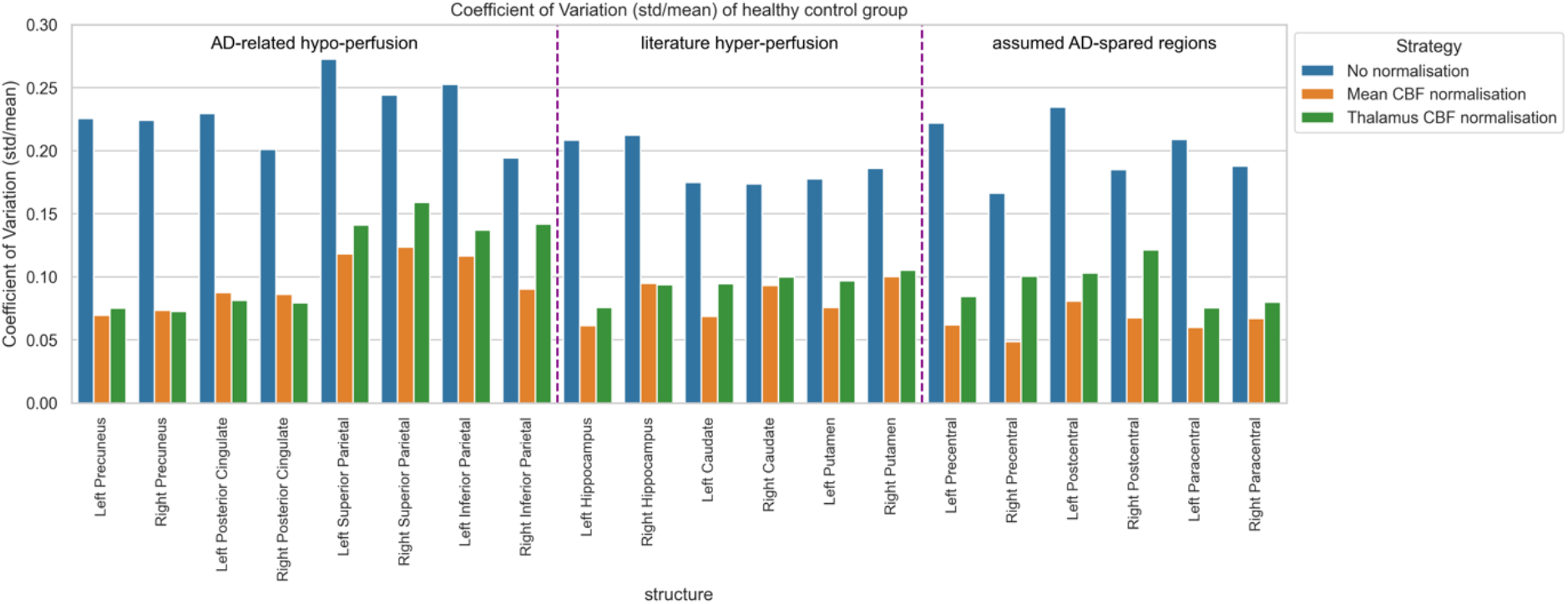
coefficients of variation of median CBF within selected ROIs averaged across healthy controls, with and without normalisation. Normalisation with mean CBF and thalamus both reduced CoV substantially.

Figure 3 shows calibrated and normalised CBF maps for one healthy subject (male between 71-75 years) and one AD subject (male between 71-75 years), respectively, chosen to be age and sex matched. Before normalisation, the AD subject was observed to have substantially lower global perfusion. After normalisation by global mean CBF, some territories of the AD subject showed higher relative perfusion than the HC (superior frontal, middle temporal and paracentral lobes shown with blue arrows). After normalisation by thalamus CBF, no territories showed higher relative perfusion on the AD subject compared to HC, and hypoperfusion in the inferior parietal regions (red arrow) was readily observed.

**Figure 3:**
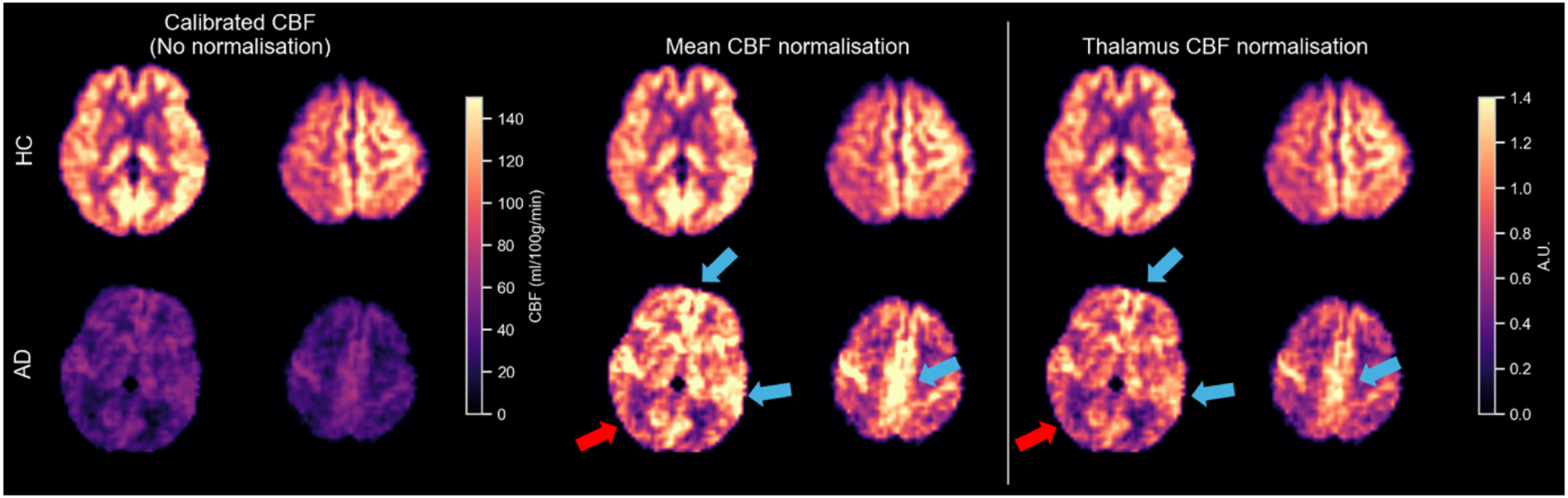
Visual comparison of CBF maps for a HC (top row) and AD subject (bottom row); without (left column) and with normalisation (central, right columns). Normalisation with mean CBF led to regions of hyperperfusion in the AD subject compared to HC (shown with blue arrows), which was not observed when normalising with thalamus CBF. Parietal lobe hypoperfusion characteristic of AD is highlighted with a red arrow.

Figure 4 shows median CBF in ROIs for all three normalisation strategies, separated into HC and AD groups. Without normalisation (top panel), AD subjects had lower CBF than HC in all ROIs. After normalisation by mean CBF, AD subjects continued to exhibit lower CBF in AD-associated ROIs, in the other two groups of ROIs the median CBF was higher in the AD group than the HC. After thalamus CBF normalisation, the median CBF in the other two ROI groups were comparable between the two cohorts, whilst the differences in AD-related ROIs were maintained.

**Figure 4:**
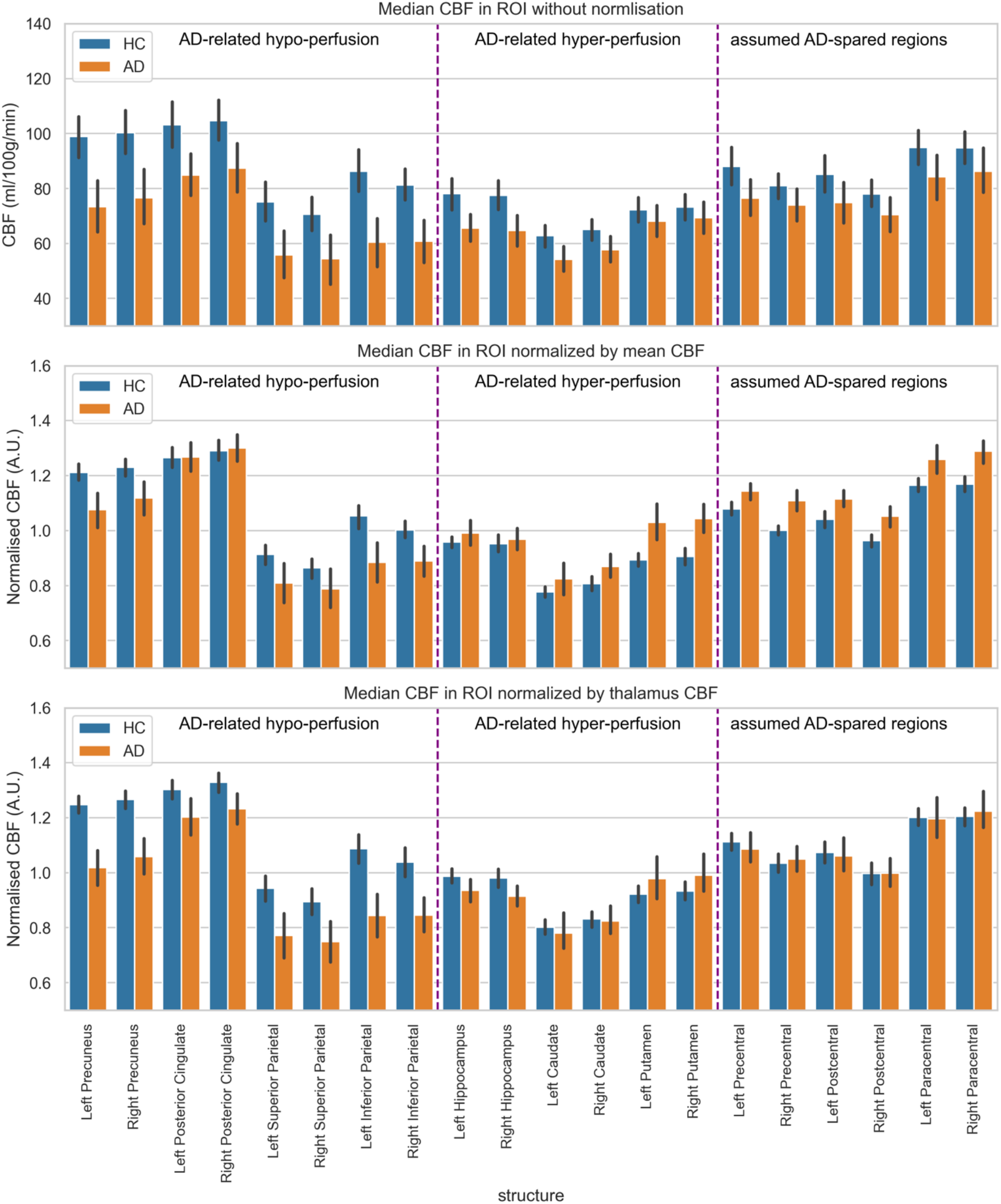
median CBF within ROIs, averaged within HC and AD groups, without normalisation (top row), normalised by mean CBF (middle row), and normalised by thalamus CBF (bottom row). Without normalisation, the AD group had lower CBF than HC in all ROIs; after normalisation by mean CBF, the AD group showed hyperperfusion in numerous ROIs, which was not the case with thalamus normalisation.

Figure 5 shows *t* statistics from a two-sampled test with the hypothesis that CBF in each ROI was higher for the HC group than AD. Only comparisons with *p* < 0.05 are shown. Compared to no normalisation, normalisation by thalamus almost always increased *t* (e.g. left and right precuneus), and sometimes resulted in statistical significance being reached where previously the comparison was not significant (e.g. right caudate). In ROIs known to be affected by AD, normalisation by mean CBF led to reduced *t* values compared to no normalisation, and in regions assumed to be spared by AD, normalisation by mean CBF revealed statistically significant comparisons with reversed sign, i.e., where AD > HC in the right paracentral lobe, for example.

**Figure 5:**
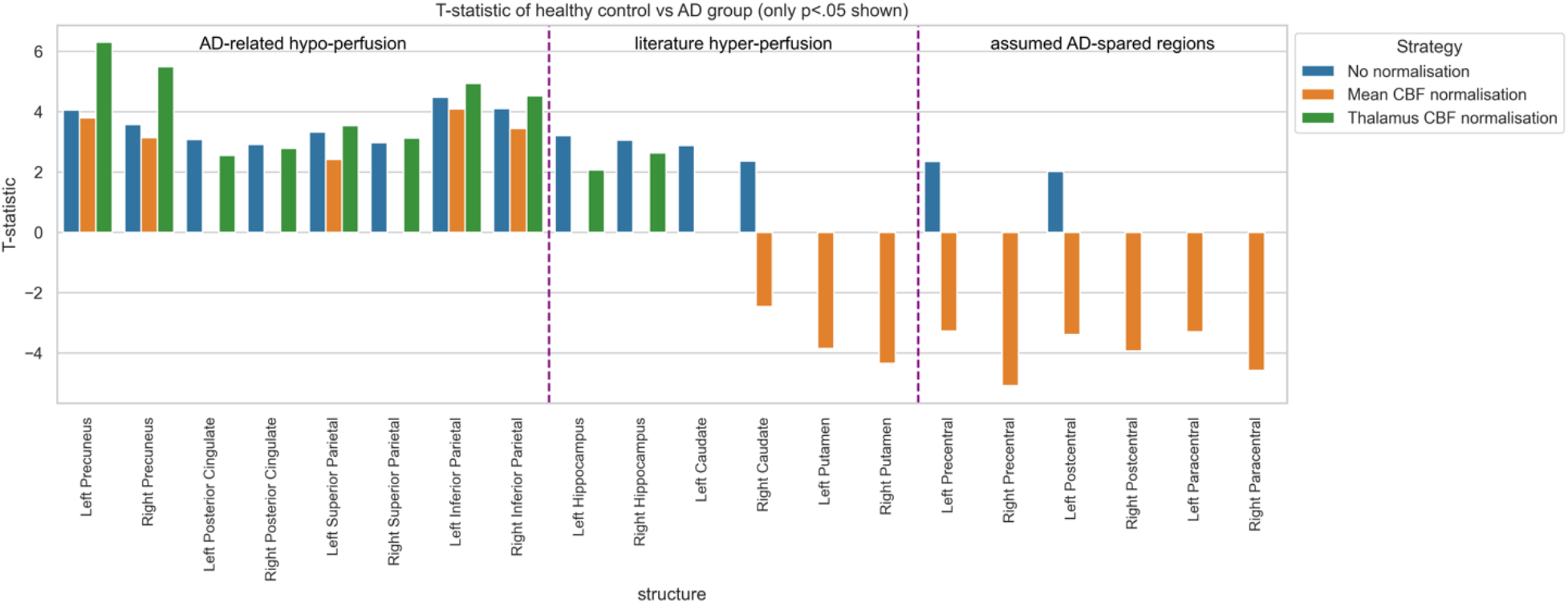
paired t-tests for the hypothesis that CBF within ROIs was higher in the HC group than AD. Only comparisons where p > 0.05 are shown. Normalisation using thalamus CBF was found to increase t-statistics in numerous ROIs compared to no normalisation. Normalisation using mean CBF was found to reverse the sign of comparisons in disease-spared regions.

## Discussion

The results presented here demonstrate the importance of choosing an appropriate normalisation strategy in the context of neurodegenerative disease. As shown in Figure 2, both normalisation strategies investigated substantially reduced the coefficient of variation of regional CBF measurements in the HC group which is desirable for group studies, however their impact on comparisons between the AD and HC group differed greatly.

A desirable property of a normalisation strategy is that it should be unaffected by disease. The results presented in Figure 1 showed that this was not the case for mean CBF, whereas thalamus CBF was more stable between the HC and AD groups. Due to the global hypoperfusion of AD subjects compared to HC, normalisation by global mean CBF (which is similar to taking a Z-transform) was found to introduce pseudo-hyperperfusion which was readily observed in Figure 3. This resulted in the emergence of statistically significant comparisons where AD perfusion exceeded HC in regions not associated with AD such as the paracentral lobes (Figure 5). These findings were in line with previously reported results for this dataset, for which normalisation by mean CBF was used. Specifically, Chen *et al*. reported hypoperfusion in the AD group compared to HC in the parietal lobes and cingulate, and hyperperfusion in postcentral gyrus and putamen [8].

The proposed alternative of normalisation using the thalamus as a reference region was found to mitigate these issues. This strategy was observed to almost always increase *t*-statistics of comparisons between the HC and AD groups in regions known to be affected by disease such as the precuneus and parietal lobes. Such a finding may be attributed to two distinct mechanisms: an increase in difference between the two groups, and/or a reduction in variance within the groups. We explored the contribution of the two mechanisms within the left precuneus (hypoperfused in the AD group) by calculating the regularised group difference as (μ_HC_ − μ_AD_)/(μ_HC_ + μ_AD_). Before and after normalisation by thalamus CBF this quantity was 0.15 and 0.10 respectively, a change of -33%. The regularised group standard deviation was calculated as 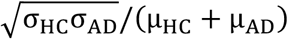; before and after normalisation this quantity was 0.13 and 0.05, a change of -61%. Although thalamus normalisation actually decreased the difference between the groups (consistent with the observation that AD subjects had global hypoperfusion, which would have been somewhat reduced by normalisation), the reduction in variance was even greater, and thus the *t*-statistic which is a function of both quantities increased: the gains from reducing variance outweighed the reduction in group difference.

A limitation of this work is that no patients with mild cognitive impairment (MCI) were included in the dataset. In MCI, global mean CBF will generally be less severely affected than for AD, and thus normalisation by global mean CBF could still be a performant strategy. This possibility would diminish as disease progresses however, reducing the overall utility of the strategy because an ideal normalisation strategy should be robust over as wide a range of disease states as possible. A further limitation of this work is the limited knowledge of how the thalamus may be affected in other dementias or indeed brain diseases: it may be a suitable choice for AD, but this does not automatically mean it will be a suitable choice for other diseases.

## Conclusion

Normalisation using mean thalamus CBF was found to be a performant strategy in the context of AD. It was found to both decrease the coefficient of variation of regional CBF measurements, and increase the detectability and magnitude of known disease effects. Compared to the commonly used strategy of normalisation by global mean CBF, the proposed strategy was found to be more robust to potential pseudo-hyperperfusion artefacts. These results demonstrate the importance that analysis decisions play in yielding statistically meaningful findings and should increase the accuracy and reliability of ASL imaging for early detection and clinical management of AD.

## Data Availability

Data may be obtained by request to the authors of this prior work: https://www.frontiersin.org/journals/neuroscience/articles/10.3389/fnins.2022.892374/full

## Acknowledgements

The authors are grateful to Hongri Chen *et al*. for providing the dataset on which this work was performed [8].

